# Interpretable Machine Learning-Based Clinical Predictive Model for Early Readmission in Patients with Cardiogenic Shock

**DOI:** 10.1101/2024.07.08.24310102

**Authors:** Xiarepati Tieliwaerdi, Abulikemu Abuduweili, Erasmus Mutabi, Kathryn Manalo, Matthew Lander

**Affiliations:** Department of Medicine, Allegheny Health Network; Robotics Institute, Carnegie Mellon University; Cardiovascular Institute, Allegheny Health Network

## Abstract

**Background/Purpose:** Cardiogenic shock (CS) is a critical condition characterized by low cardiac output leading to end-organ hypoperfusion and often multisystem organ failure, affecting up to 50,000 people annually in the United States. Acute myocardial infarction (AMI) is the primary cause, responsible for 81% of CS cases. Despite advancements in reperfusion therapies improving survival, in-hospital mortality remains high at 40%-67%, with 18.6% of survivors readmitted within 30 days. Traditional methods struggle to quantify and process the complex interactions among various risk factors, making prediction of readmissions challenging. Machine learning (ML) offers a promising solution by capturing intricate patterns and non-linear relationships among numerous variables. This study aims to develop an ML-based prediction model for 7-day and 30-day readmission rates in CS patients using the 2019 National Readmission Database (NRD). Additionally, the study utilizes SHapley Additive exPlanations (SHAP) to interpret the outcomes of the applied machine learning methods.

**Method:** We conducted a retrospective study using the NRD for 2019. Index hospitalizations were identified by non-elective admissions with a primary ICD-10 diagnosis of cardiogenic shock. Exclusions included patients under 18, missing length of stay or days to event data, and same-day transfers. The primary outcome was readmission within 7- and 30-days post-discharge. Welch’s t-test compared continuous variables. Various ML models were evaluated for their predictive performance, and SHAP values were used to interpret the most influential features.

**Results:** The study included 97,653 adults hospitalized for CS, with a mean age of 65.8 years and 38.4% being female. The in-hospital mortality rate was 33.7%. Among 51,976 index hospitalizations, 8.3% were readmitted within 7 days, and 21.02% within 30 days. Significant predictors of higher readmission rates included younger age, lower income, Medicaid insurance, CKD3, drug abuse, chronic pulmonary disease, PHTN, depression, leukemia, lymphoma, discharge against medical advice, and certain hospital characteristics. The FT-Transformer (a specialized deep neural network approach for tabular data) model achieved the highest AUCs of 0.76 and 0.78 for 7-day and 30-day readmissions, respectively, outperforming traditional methods like Logistic Regression (AUCs: 0.60 and 0.63).

SHAP analysis revealed a wide array of features contributing to readmission predictions at both the population and individual levels. For the general population, the top features included APRDRG, DRG_NoPOA, age, chronic kidney disease, length of stay, number of ICD-10 codes, and disposition at discharge. In contrast, for an individual patient, the most influential feature for predicting 7-day readmission may differ, though there are some overlaps. This highlights the potential of personalized medicine, where individual risk factors are weighted differently compared to the general population, providing tailored insights for targeted interventions.

**Conclusion:** This study demonstrates that advanced ML models, particularly the FT-Transformer and Random Forest, significantly outperform traditional methods in predicting readmissions in CS patients. The use of SHAP values enhances the interpretability of these models, providing actionable insights for healthcare providers. The differentiation between general population feature contributions and individual-specific factors underscores the importance of personalized medicine. By understanding individual risk profiles, healthcare providers can implement more precise and effective interventions, ultimately aiming to reduce readmissions and optimize healthcare outcomes.

## 1 Introduction

Cardiogenic shock (CS) is a low-output cardiac state with a high-acuity, potentially complex, and hemodynamically diverse state of end-organ hypoperfusion that is frequently associated with multisystem organ failure [1]. It affects up to 50,000 people annually in the United States, with acute myocardial infarction (AMI) as the most frequent cause accounting for 81% of all the CS cases [23]. Over the years, tremendous advances in reperfusion therapies have improved survival. However, in-hospital mortality remains high ranging from 40%-67% and so do re-admission rates [45]. Among patients who survive cardiogenic shock following AMI, about 18.6% get re-admitted within 30 days 6]. Predictors of re-admission include low economic status, female sex, atrial fibrillation, ventricular tachycardia, and mechanical circulatory device placement. Congestive heart failure and new myocardial infarction (MI) are the most common causes of re-admission [7,8,9]. Although those risk factors have been well recognized, physicians still face significant challenges in timely quantifying and processing complex interactions among various risk factors. In addition, the challenge lies in predicting their combined impact on ultimate healthcare outcomes as they present in diverse combinations across different individuals.

The advent of machine learning (ML) in healthcare offers a potential solution to these challenges. When compared to traditional predictive methods, it is better at capturing intricate interactions and patterns as well as non-linear relationships among a vast number of variables [10,11]. It holds potential in developing clinical prediction models that are reliable and cost-effective. It has wide medical applications ranging from health records management to interpreting imaging, research, and personalized medicine [12]. Multiple studies have proposed ML based prediction models as clinical decision support tools in CS management and achieved good results [13,14,15]. Golas *et al.* used deep neural network to develop a risk prediction model for 30-day re-admissions in heart failure patients, which performed better than the traditional methods [16]. Inspired by the success of the Transformer neural network across a range of AI applications [17], including computer vision, natural language processing, and robotics, Yuri et al. introduced the FT-Transformer (Feature-tokenizer Transformer) neural network specifically for tabular data prediction [18]. This model has demonstrated superior performance over previous methods on most tabular data prediction tasks. Given that our readmission prediction challenge also falls within the realm of tabular data prediction, we selected the FT-Transformer as our base model for this study.

Deep neural network algorithms have the potential to outperform traditional methods. However, their lack of interpretability has been a significant obstacle to their application in the medical field. Providers need to understand how a prediction model derives its conclusions, especially in high-stakes areas such as medicine. To address this challenge, SHapley Additive exPlanations (SHAP) method was proposed to interpret the any machine learning prediction results [19], including deep neural networks.

The national re-admission database is a powerful and unique database designed to support various analyses of national re-admissions for all patients, including matrices such as re-admission rates by diagnosis, procedure, patient demographics, or expected payment source. Given its volume and richness in data, we are using the national re-admission database from 2019 to develop a prediction model via machine learning algorithms for 7-day and 30-day re-admission rates in patients with CS. To our knowledge, prior studies have yet to be done using this database to develop such a prediction model. This study seeks to provide predictions of 7-day and 30-day re-admissions and the most likely causes. This study employs SHAP method to enhance interpretability of predictions, enabling healthcare providers to make informed decision and implement targeted interventions to reduce re-admissions and optimize healthcare outcomes.

## 2 Method

### Design

The objective of this study was to develop a machine learning model that predicts the probability of patient readmission within 7 and 30 days, based on basic patient information. In addition to striving for accurate predictions, special attention is given to interpreting the results of the machine learning models.

### Data

We performed a retrospective study using the Agency for Health-care Research and Quality Health-care Cost and Utilization Project, Nationwide Readmissions Database (NRD) for the year 2019. CS index hospitalizations were defined as non-elective admission with a primary International Classification of Diseases and Related Health Problems (ICD)-10 diagnosis code of cardiogenic shock (R57.0). Index hospitalizations were excluded if: 1) The patients were younger than 18 years; 2) there was no information on the length of stay (LOS) or Daystoevent. 3) transfer or same-day stay involving multiple discharges (aka :SAMEDAYVENT not equals 0). Variable “NRD_visitlink” was used to identify the patients and the time between the two admissions was obtained by subtracting the variable “NRD_DaysToEvent.” Subtracting length of stay of index admissions from time between two admissions provided the interval time to readmission. Index hospitalizations were studied between January to November to facilitate identification of 7-d and 30-d readmission rates for all discharged patients for the 2019 calendar year. For index hospitalizations with more than one readmission within 30 d, only the initial admission for calendar year per patient was included for analysis as an index admission. Welch’s t test was used to compare continuous variables. The primary outcome was defined as CS readmission that occurred within the first 7 days and 30 days of discharge from last CS index hospitalization.

### Settings

In this study, ICD-10 (International Classification of Diseases, Tenth Revision) codes were truncated to three figures to reduce the dimensionality of our data. For example, the original code ’I5023’ in NRD database was shortened to ’I50’, allowing us to focus on general conditions such as “’I50 - heart failure” instead of more specific diseases. This adjustment decreased the number of ICD-10 feature dimensions from 3,687 to 1,182. Reducing the feature dimensionality, which results in more patient cases being categorized under each general disease category, enables the machine learning models to acquire broader knowledge. This approach minimizes the risk of overfitting on specific features, promoting a more generalized model performance.

In our machine learning experiments, the patient population was divided into three distinct sets: 60% for model training, 20% for validation, and 20% for testing. The models were trained on the training set, with the validation set used for parameter tuning. After training, the performance of the models was evaluated on the test data using the AUC-ROC (Area Under the Receiver Operating Characteristic Curve) metric.

### Prediction Model

In this study, we utilized FT-Transformer [18] as our base prediction model. This model integrates the Feature Tokenizer (FT), which converts input numerical and categorical features—such as age and diagnosis of heart diseases—into vector embeddings. Subsequently, the Transformer neural network, a novel architecture with attention mechanisms, is employed to predict the probability of readmission. For comparative purposes, we also conducted experiments using a range of established methods, including Random Forest, SVM (Support Vector Machine), XGBoost (Extreme Gradient Boosting), LightGBM (Light Gradient Boosting Machine), Decision Tree, AdaBoost (Adaptive Boosting), and Logistic Regression.

### Interpretation of outcome

Understanding the rationale behind a model’s prediction is often as crucial as the accuracy of the prediction itself, especially in medical applications. However, achieving the highest accuracy on large modern datasets frequently involves complex models that are challenging to interpret, such as ensemble or deep learning models. In this study, we employed SHAP (SHapley Additive exPlanations) [19] to analyze feature contributions and provide interpretability to the models’ predictions. SHAP assigns an importance value to each feature for a specific prediction, facilitating a deeper understanding of the decision-making process within any prediction model.

## 3 Results

### Baseline characteristics

The study included 97,653 adults who was hospitalized for CS. 38.4% of those patients were female. The mean age was 65.8 years. 32,881 of them died during their presentation with a mortality rate of 33.7%. Index hospitalizations were 51,976; 7-day readmission was 4317, 7-day readmission rate was 8.3%; 30-day readmission was 10,927, 30 day readmission rate was 21.02%.

Table 1 summarizes the baseline characteristics of populations during index hospitalization. We reported only the 10 statistically significant factors in Table 1. For a full version of the baseline characteristics, please refer to Table A. 1 in the appendix. A p-value of <0.05 is considered statistically significant, while a p-value of <0.001 is considered highly significant. Table 1 reveals that factors significantly associated with higher 7-day and 30-day readmission rates include younger age, lower income, Medicaid insurance, drug abuse, chronic pulmonary disease, diabetes mellitus with complications, hypertension, and discharge against medical advice. Conversely, factors such as private insurance and routine discharge are associated with lower readmission rates.

**Table 1.** Baseline Characteristics of Study Population (10 Statistically Significant Factors)

| Characteristics | Index Admission (%) | 7-day readmission rate (%) |  |  | 30-day readmission rate (%) |  |  |
| --- | --- | --- | --- | --- | --- | --- | --- |
|  |  | readmission | no readmission | p value | readmission | no readmission | p value |
| Mean age | 65.90 | 65.11 | 65.97 | <0.001 | 65.27 | 66.06 | <0.001 |
| Lowest income | 28.80 | 30.00 | 28.60 | 0.050 | 30.70 | 28.20 | <0.001 |
| Medicaid | 13.70 | 16.80 | 13.40 | <0.001 | 16.50 | 13.00 | <0.001 |
| Private insurance | 19.20 | 16.70 | 19.40 | <0.001 | 15.80 | 20.10 | <0.001 |
| Drug abuse | 3.10 | 4.60 | 3.00 | <0.001 | 4.20 | 2.80 | <0.001 |
| Chronic pulmonary disease | 27.30 | 30.30 | 27.10 | <0.001 | 30.40 | 26.50 | <0.001 |
| Diabetes mellitus with complications | 3.40 | 4.90 | 3.30 | <0.001 | 4.50 | 3.10 | <0.001 |
| Hypertension | 11.80 | 8.50 | 12.10 | <0.001 | 7.60 | 12.90 | <0.001 |
| Routine | 33.60 | 29.40 | 34.00 | <0.001 | 29.80 | 34.60 | <0.001 |
| Against medical advice | 1.30 | 3.00 | 1.10 | <0.001 | 2.40 | 1.00 | <0.001 |

### Prediction performance

Figure 1 presents the ROC curves and ROC-AUC scores of various machine learning methods for predicting 7-day and 30-day readmissions. The FT-Transformer consistently outperformed other models, achieving AUCs of 0.76 and 0.78, respectively. Random Forest also demonstrated strong performance with AUCs of 0.71 and 0.76. SVM showed moderate effectiveness with AUCs of 0.73 and 0.70, as did XGBoost with AUCs of 0.68 and 0.73, and LightGBM with AUCs of 0.66 and 0.72. Decision Tree, Logistic Regression, and AdaBoost had the lowest AUCs, ranging from 0.60 to 0.64, indicating less reliable performance. Notably, Logistic Regression, a traditional analytical method, had AUCs of 0.60 and 0.63, significantly lower than FT-Transformer and Random Forest. These results suggest that the FT-Transformer is the most effective model for predicting readmissions in this patient population, offering valuable insights for targeted interventions to reduce readmission rates. It is important to note that 7-day readmission is a more class-imbalanced problem, with far fewer positive samples (7-day readmissions) compared to negative samples (no 7-day readmissions). Random Forest’s performance was significantly impacted by this issue, with a 9% reduction in AUC (from 0.76 to 0.70) when predicting 7-day readmission compared to 30-day readmission. However, the FT-Transformer’s performance drop was much smaller, at about 3%. This comparison highlights the superior predictive capabilities of advanced machine learning models, such as FT-Transformer, over traditional methods like Logistic Regression.

**Figure 1.**
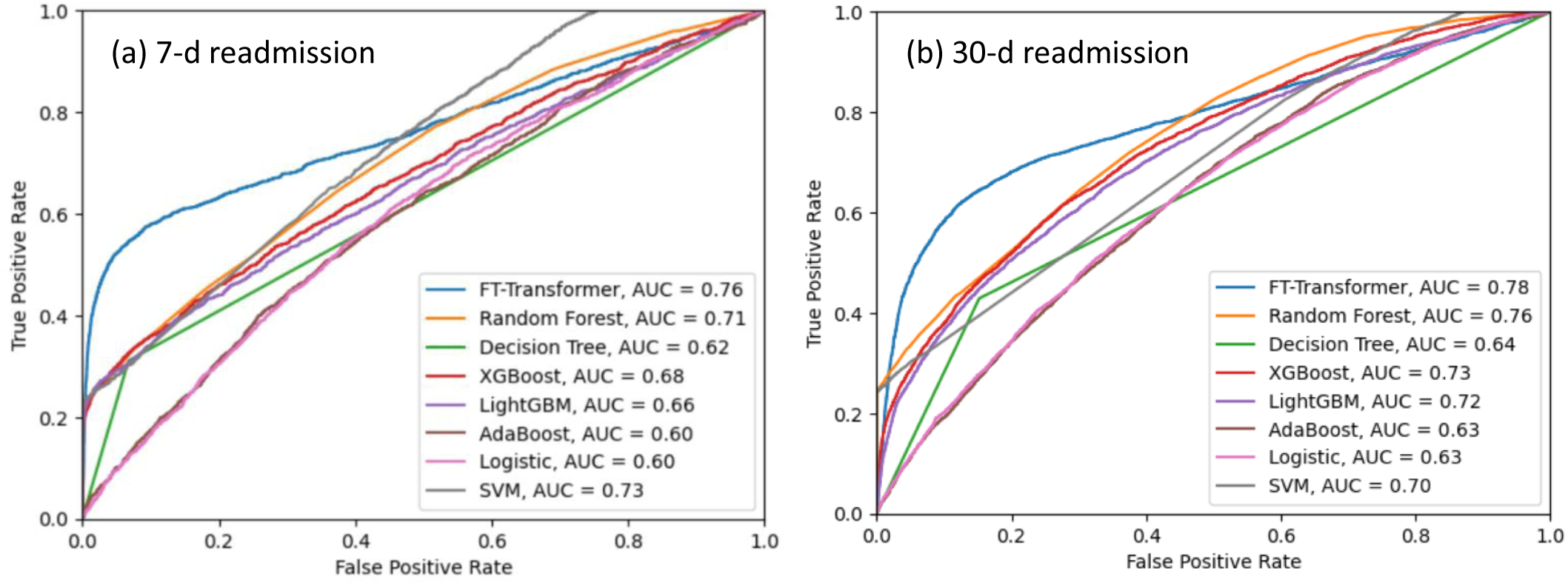
ROC Curves for Different ML Algorithms. This figure compares the performance of various machine learning algorithms in predicting (a) 7-day readmission and (b) 30-day readmission.

### Feature importance

Figure 2 demonstrates the contributions of top 10 most influential features to predicting 7-day readmission in patients with cardiogenic shock using SHAP values from a FT-transformer model. SHAP value denotes the importance of each feature. Notably, no single feature dominates the prediction. The top 10 features, including APRDRG, DRG_NoPOA, age, chronic kidney disease, length of stay, number of ICD-10 codes, disposition at discharge, essential hypertension, other medical care, and COPD, each contribute SHAP values between 0.01 and 0.03. Meanwhile, the cumulative contribution of the remaining 1190 features is 0.39. This distribution indicates that the prediction of cardiogenic shock readmission is highly complex and multidimensional, relying on a wide array of factors rather than a few predominant ones. Understanding this complexity can help healthcare providers better target interventions to reduce readmission rates.

**Figure 2.**
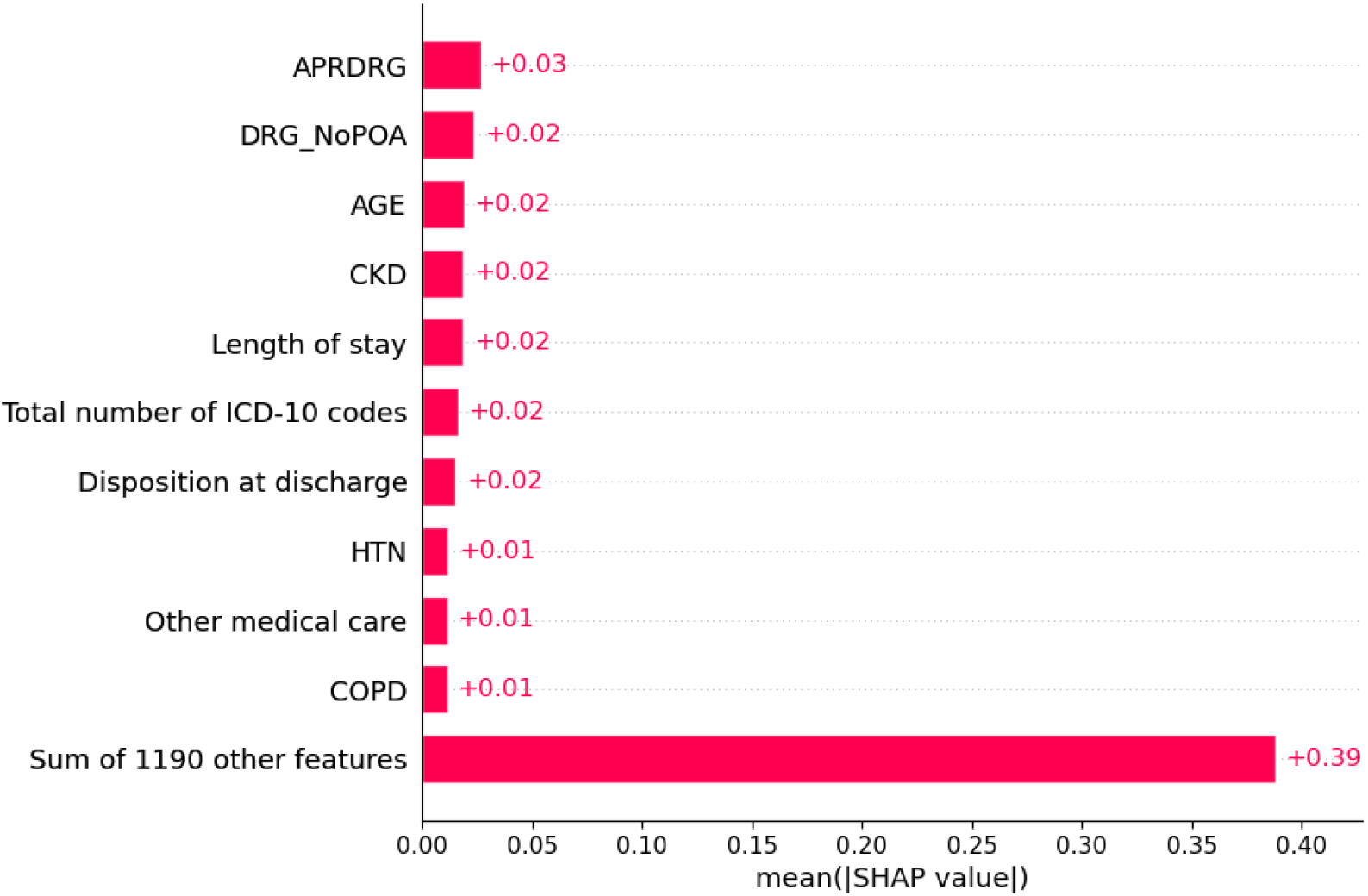
Feature contributions to predicting 7-day readmission in study population. APRDRG: All patient refined diagnosis related groups, which is an index of case severity and complexity used for insurance reimbursement; DRG_NoPOA: Diagnosis related groups assignment made without the use of the present on admission flags for the diagnoses; CKD: Chronic kidney disease; HTN: Essential hypertension; Other medical care: including palliative care, blood transfusion, chemotherapy or radiation therapy for neoplasm or other unspecified medical care; COPD: chronic obstructive pulmonary disease and heart failure.

Similarly, Figure 3 demonstrates the contributions of the top 10 most influential features for predicting 30-day readmissions in patients. Several key features overlap with those identified for 7-day readmission predictions, such as DRG_NoPOA, length of stay, APRDRG, number of ICD-10 codes, and disposition at discharge. However, unique to the 30-day prediction are significant features such as DNR status and heart failure. Despite these differences, no single feature overwhelmingly dictates the prediction outcomes, similar to the 7-day predictions. The top 10 features contribute SHAP values ranging from 0.02 to 0.04, while the cumulative contribution of the remaining 1,190 features totals 0.69.

**Figure 3.**
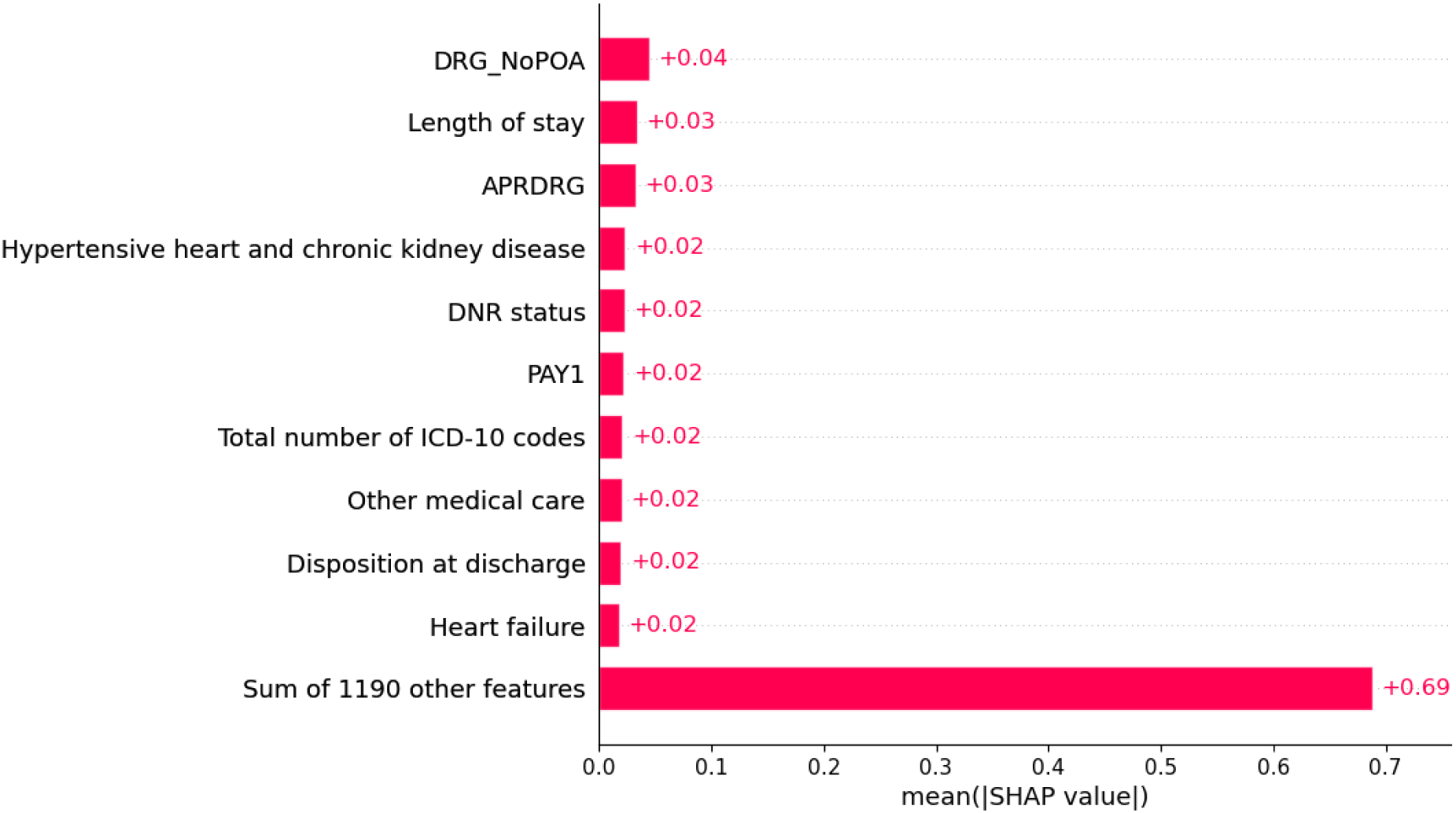
Feature Contributions to Predicting 30-Day Readmission.

### Interpretation of outcome

Figure 2 and Figure 3 illustrate the overall importance of features across all datasets. However, statistical averages may not always be relevant to individual cases. A key aspect of our study is the ability to interpret specific prediction outcomes for individual cases. We utilize bar plots of SHAP value to display the impact of each feature on the prediction outcome, providing a clear and direct visualization of how various factors contribute to the model’s predictions.

Figure 4 presents the interpretation of FT-Transformer model’s 7-day readmission prediction for an individual patient, where the true label is positive, indicating an expected readmission within 7 days. The predicted readmission probability by the FT-Transformer is p=0.99, confirming that the model accurately anticipated the readmission. In our visualization, red bars (positive) indicate features that increase the readmission probability, whereas blue bars (negative) indicate features that decrease it. Notably, the presence of portal vein thrombosis, marked as ’1= portal vein thrombosis’ in the figure (where 1 indicates positive), significantly increases the readmission probability by 37%. Additionally, an APRDRG value of 11 increases the probability by 7%. Other contributing factors include COPD, which increases the likelihood by 5%, and conditions like atrioventricular and left bundle-branch block (AV and LBBB) and hypertension (HTN). In contrast, dysphagia slightly reduces the readmission probability by 2%. The collective impact of the remaining 1,191 features accounts for a 22% increase, illustrating that while they contribute to the overall prediction, portal vein thrombosis remains the dominant factor for this case. It is important to note that the contributions of individual features are not isolated but are interconnected, resulting in a combined effect on the overall prediction.

**Figure 4.**
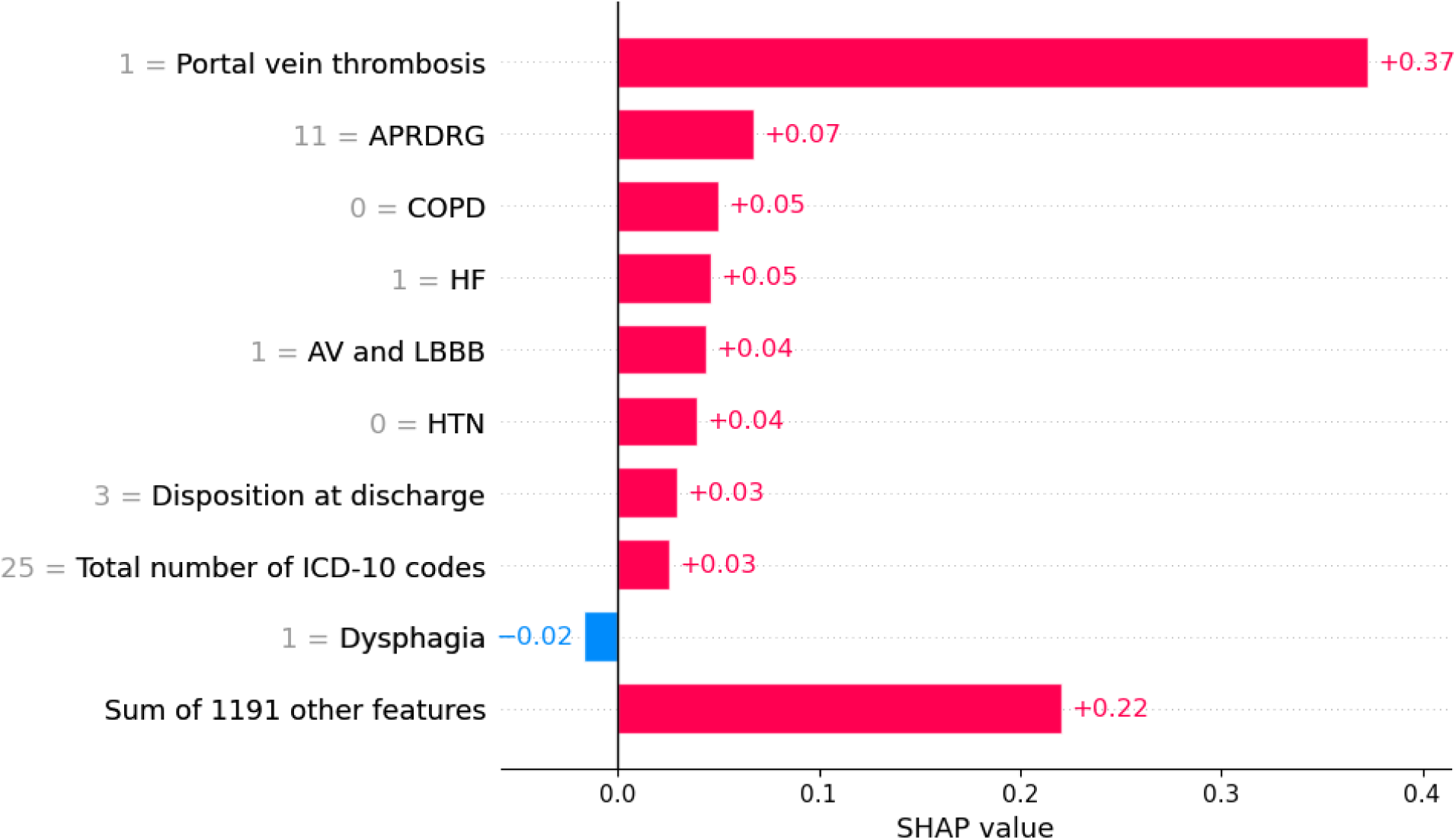
Feature Contributions for Predicting 7-Day Readmission. This figure displays the feature contributions for a specific individual who was readmitted within 7 days. The model predicted the readmission with a high probability of 0.99.

Similarly, Figure 5 presents the interpretation of FT-Transformer model’s 30-day readmission prediction for an individual patient, where the true label is positive, indicating an expected readmission within 30 days. The predicted readmission probability by the FT-Transformer is p=0.99, confirming that the model accurately anticipated the readmission. Notably, the presence of Drainage, significantly increases the readmission probability by 16%. Additionally, an Pleural effusion increases the probability by 10%. Other contributing factors include COPD, which increases the likelihood by 6%. In contrast, a disposition at discharge coded as ’4’ slightly decreases the probability by 5%. Collectively, the impact of the remaining 1,191 features accounts for a 25% increase in readmission probability. n the appendix, additional interpretation results are provided, as depicted in Figure A. 1 and Figure A. 2.

**Figure 5.**
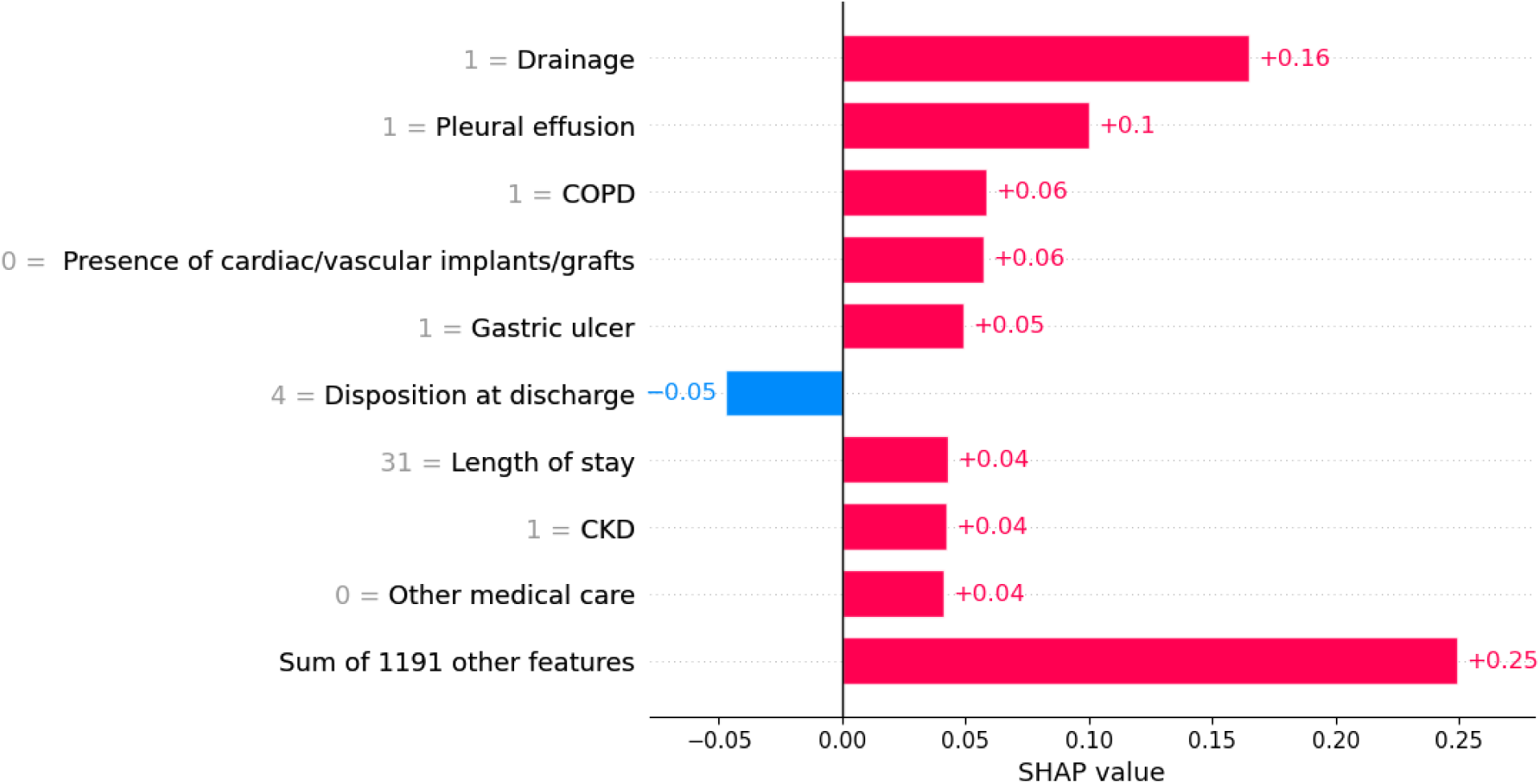
Feature Contributions for Predicting 30-Day Readmission. This figure displays the feature contributions for a specific individual who was readmitted within 30 days. The model predicted the readmission with a high probability of 0.99.

## 4 Discussion

### Statistical Study

This retrospective study utilizing the 2019 Nationwide Readmissions Database (NRD) provides valuable insights into the readmission rates of patients with cardiogenic shock (CS). Our analysis included 97,653 adults, revealing a mean age of 65.8 years and a 33.7% mortality rate during initial hospitalization. Index hospitalizations totaled 51,976, with 7-day and 30-day readmission rates of 8.3% and 21.02%, respectively. Key factors associated with higher readmission rates include younger age, lower income, Medicaid insurance, chronic kidney disease (CKD3), drug abuse, chronic pulmonary disease, pulmonary hypertension (PHTN), depression, leukemia, lymphoma, and discharge against medical advice. Conversely, private insurance and routine discharge were linked to lower readmission rates.

### Machine Learning Study

Machine learning models were employed to predict readmission rates, with the FT-Transformer demonstrating superior performance (AUCs: 0.76 and 0.78) compared to other methods. In contrast, traditional methods like Logistic Regression had significantly lower AUCs (0.60 and 0.63). The advanced machine learning models outperformed Logistic Regression by providing better discrimination between readmitted and non-readmitted patients. Logistic Regression, while useful for understanding linear relationships and providing interpretable coefficients, falls short in capturing the complex, nonlinear interactions that machine learning models can identify. This underscores the enhanced predictive capabilities of advanced machine learning techniques, which are particularly valuable in high-dimensional datasets common in healthcare

### Interpretation of machine learning models

SHAP values provide a means to interpret the contributions of various features in a machine learning model’s prediction. At the population level, SHAP values are averaged across all instances (patients) in the dataset. This approach helps identify features that consistently contribute to predictions across the entire patient population. In our study, the population-level SHAP analysis did not reveal a single dominating predictor. Instead, it showed that multiple features such as APRDRG -an index of case severity and complexity, DRG_NoPOA, which identifies diagnoses not present at admission and assesses hospital-acquired conditions, age, chronic kidney disease, and length of stay, each contributed modestly to the prediction of 7-day readmission, with individual SHAP values ranging from 0.01 to 0.03, as shown in Figure 2. At the individual level, SHAP values reflect the contribution of each feature to the prediction for a specific patient. This can result in a single feature having a dominant influence on the model’s prediction. For instance, in one patient’s case as shown in Figure 4, portal vein thrombosis had a SHAP value of +0.37, making it the dominant predictor for that individual’s readmission risk. The discrepancy between SHAP values at the population level and individual level highlights the complexity and variability of predictive factors in patients with cardiogenic shock. While general trends are valuable for understanding broad risk factors, individual predictions enable tailored interventions that address specific patient needs with goal of personalized medicine. This type of personalized analysis has the potential to reduce readmission rates. For instance, the interpretation provided indicates that portal vein thrombosis is a key contributor to the readmission risk for the patient shown in Figure 4, it’s crucial to note that these interpretations, derived from mathematical calculations, should serve as references for healthcare providers to consider if targeted interventions could effectively reduce readmission risks.

## 5 Conclusion

This study shows the complexity of predicting 7-day and 30-day readmission rates in patients with cardiogenic shock. Advanced machine learning models like FT-Transformer outperformed traditional machine learning methods, capturing complex, nonlinear interactions for more accurate predictions. SHAP value analysis revealed interpretation of contributing features in a study population level as well as individual level. These findings can help healthcare providers target interventions to reduce readmission rates, highlighting the potential of integrating advanced machine learning into clinical practice for better patient outcomes and resource optimization.

## Data Availability

All data produced in the present study are available upon reasonable request to the authors.

## Appendix

### Baseline characteristics

Table A. 1 summarizes the baseline characteristics of populations during index hospitalization.

**Table A.1.** Baseline Characteristics of Study Population.

| Characteristics | Index Admission (%) | 7-day readmission rate (%) |  |  | 30-day readmission rate (%) |  |  |
| --- | --- | --- | --- | --- | --- | --- | --- |
|  |  | readmission | No readmission | p value | readmission | No readmission | p value |
| Female | 36.10 | 35.90 | 36.10 | 0.741 | 36.70 | 35.90 | 0.133 |
| Mean age | 65.90 | 65.11 | 65.97 | <0.001 | 65.27 | 66.06 | <0.001 |
| Lowest income | 28.80 | 30.00 | 28.60 | 0.050 | 30.70 | 28.20 | <0.001 |
| Highest income | 19.60 | 18.70 | 19.70 | 0.091 | 18.30 | 20.00 | <0.001 |
| Medicare | 61.30 | 61.60 | 61.30 | 0.683 | 63.20 | 60.80 | <0.001 |
| Medicaid | 13.70 | 16.80 | 13.40 | <0.001 | 16.50 | 13.00 | <0.001 |
| Private insurance | 19.20 | 16.70 | 19.40 | <0.001 | 15.80 | 20.10 | <0.001 |
| Self-pay | 2.50 | 2.00 | 2.50 | 0.031 | 1.90 | 2.60 | <0.001 |
| CKD3 | 16.20 | 17.70 | 16.10 | 0.008 | 18.60 | 15.60 | <0.001 |
| CKD4 | 4.80 | 5.00 | 4.80 | 0.384 | 5.30 | 4.60 | 0.003 |
| CKD5 | 0.50 | 0.60 | 0.40 | 0.050 | 0.50 | 0.40 | 0.143 |
| Unspecified HF | 4.20 | 4.20 | 4.20 | 0.969 | 4.00 | 4.20 | 0.219 |
| Right HF | 1.10 | 1.00 | 1.10 | 0.465 | 1.00 | 1.10 | 0.577 |
| Chronic systolic HF | 7.80 | 7.60 | 7.80 | 0.595 | 7.60 | 7.80 | 0.413 |
| Heart transplant | 0.10 | 0.10 | 0.10 | 0.691 | 0.10 | 0.00 | 0.075 |
| Acute MI | 30.50 | 29.90 | 30.50 | 0.352 | 26.90 | 31.40 | <0.001 |
| LVAD Present | 0.80 | 0.90 | 0.80 | 0.753 | 1.00 | 0.80 | 0.017 |
| Drug abuse | 3.10 | 4.60 | 3.00 | <0.001 | 4.20 | 2.80 | <0.001 |
| Chronic pulmonary disease | 27.30 | 30.30 | 27.10 | <0.001 | 30.40 | 26.50 | <0.001 |
| PHTN | 16.50 | 16.60 | 16.50 | 0.869 | 17.90 | 16.10 | <0.001 |
| Dementia | 3.40 | 3.30 | 3.50 | 0.566 | 3.20 | 3.50 | 0.155 |
| Peripheral Vascular Disease | 4.00 | 4.60 | 4.00 | 0.050 | 4.60 | 3.80 | <0.001 |
| BMI >35 | 10.50 | 10.40 | 10.50 | 0.802 | 10.70 | 10.40 | 0.383 |
| Diabetes mellitus with complications | 3.40 | 4.90 | 3.30 | <0.001 | 4.50 | 3.10 | <0.001 |
| Hypertension | 11.80 | 8.50 | 12.10 | <0.001 | 7.60 | 12.90 | <0.001 |
| Cerebral infarction | 3.20 | 3.20 | 3.20 | 0.919 | 3.20 | 3.20 | 0.903 |
| Cerebral hemorrhage | 0.70 | 0.50 | 0.80 | 0.074 | 0.60 | 0.80 | 0.060 |
| Depression | 8.80 | 9.40 | 8.80 | 0.168 | 9.50 | 8.70 | 0.005 |
| Leukemia | 1.20 | 1.50 | 1.20 | 0.093 | 1.50 | 1.10 | 0.004 |
| Lymphoma | 1.90 | 2.20 | 1.80 | 0.103 | 2.20 | 1.80 | 0.004 |
| Solid tumor | 1.40 | 1.20 | 1.40 | 0.385 | 1.40 | 1.30 | 0.969 |
| Metastatic cancer | 1.90 | 1.80 | 1.90 | 0.534 | 2.10 | 1.80 | 0.129 |
| Small bed size | 10.90 | 11.60 | 10.90 | 0.154 | 10.40 | 11.10 | 0.050 |
| Medium bed size | 23.60 | 23.90 | 23.50 | 0.595 | 23.20 | 23.70 | 0.263 |
| Large bed size | 65.50 | 64.50 | 65.60 | 0.158 | 66.40 | 65.20 | 0.022 |
| Metropolitan teaching | 81.50 | 81.90 | 81.50 | 0.552 | 82.30 | 81.30 | 0.021 |
| Metropolitan non-teaching | 15.20 | 14.90 | 15.20 | 0.685 | 14.80 | 15.30 | 0.210 |
| Non-metropolitan | 3.30 | 3.20 | 3.30 | 0.633 | 2.90 | 3.40 | 0.013 |
| Routine | 33.60 | 29.40 | 34.00 | <0.001 | 29.80 | 34.60 | <0.001 |
| Transfer to facility | 33.00 | 33.60 | 32.90 | 0.374 | 35.30 | 32.30 | <0.001 |
| Home health care | 27.40 | 27.00 | 27.40 | 0.518 | 28.10 | 27.20 | 0.075 |
| Against medical advice | 1.30 | 3.00 | 1.10 | <0.001 | 2.40 | 1.00 | <0.001 |

### Interpretation of outcome

In Figure 4 and Figure 5, we interpret the outcomes of the prediction model using bar plots of SHAP values. In this section, we illustrate the outcomes using a waterfall plot of SHAP values. Figure A. 1 presents the interpretation of the FT-Transformer model’s 30-day readmission prediction with a waterfall plot of SHAP values, where the true label is positive, indicating an expected readmission within 30 days. The predicted readmission probability by the FT-Transformer is p=0.988, confirming that the model accurately anticipated the readmission. The figure shows that the base predicted readmission rate is 0.234, *E*[*f*(*x*)] = 0.234, derived from the statistics of the training data, indicating that approximately 23.4% of cases in the training set were readmitted. The feature contributions are as follows: 1,191 other features collectively increase the readmission probability by 12%. Additionally, certain early complications of trauma and the NCHS urban-rural classification scheme increase the likelihood by 3% and 5%, respectively. The length of stay of 31 days increases the likelihood by 7%, while being classified under PCLASS_ORPRC as ’2’ increases it by 8%. Disorders of pancreatic internal secretion increase it by 8%. Unspecified diseases of blood and blood-forming organs decrease the likelihood by 10%. Drainage increases it by 10%, APRDRG coded as ’22’ increases it by 11%, and DNR status, as the most significant factor, increases the likelihood by 21%. These contributions combine to yield a predicted probability of 0.988, *f*(*x*) = 0.988.

**Figure A. 1.**
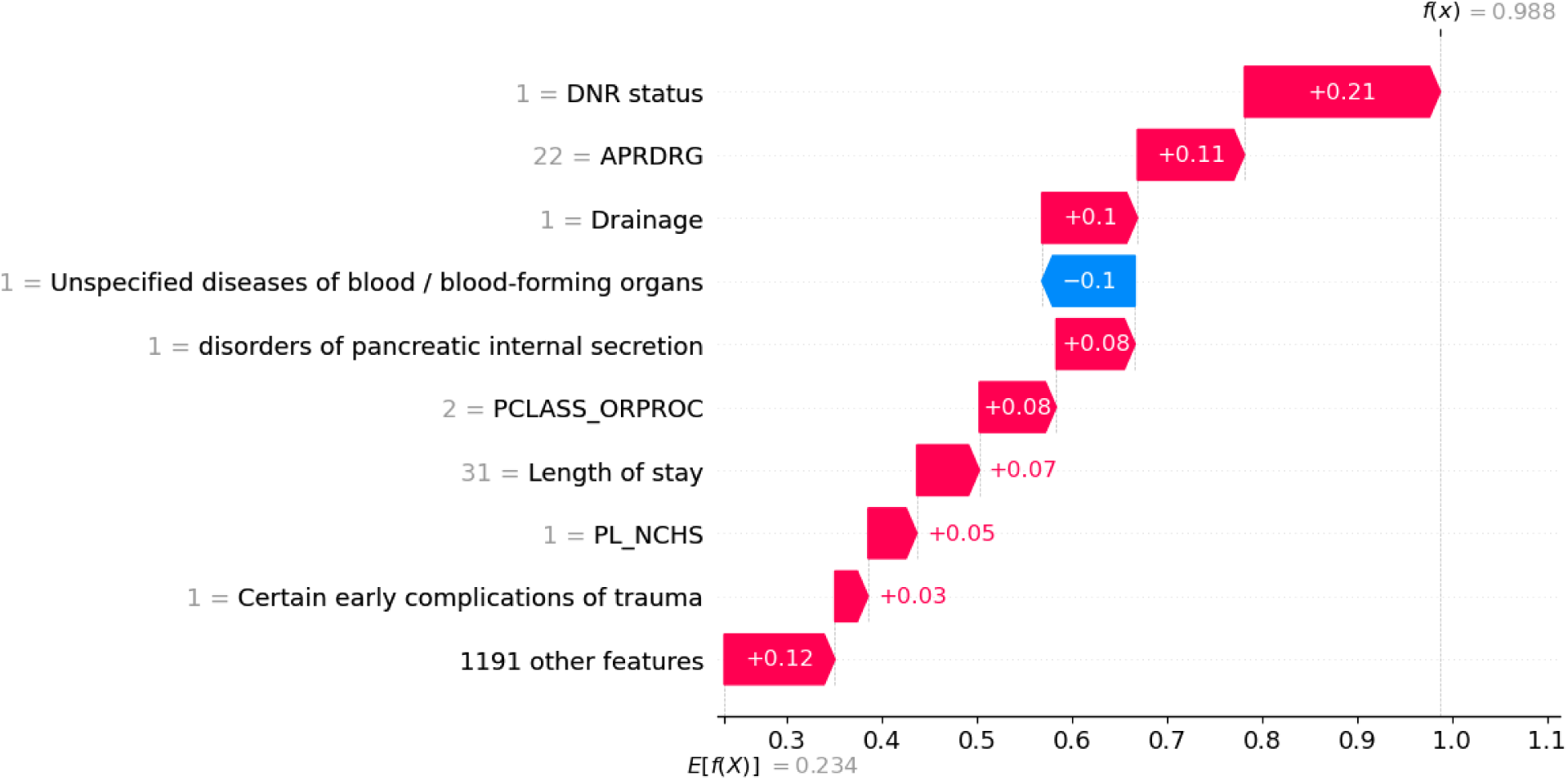
Feature Contributions for Predicting 30-Day Readmission. This figure displays the feature contributions for a specific individual who was readmitted within 30 days. The model predicted the readmission with a high probability of 0.988.

Figure A. 2 presents the interpretation of a negative sample with a waterfall plot of SHAP values. This figure illustrates the prediction model’s interpretation for a case with a true negative label, indicating no readmission within 30 days. The FT-Transformer model predicted a readmission probability of p=0.006, accurately forecasting the lack of readmission. Mirroring the approach in Figure A. 1, the base expected readmission rate is 0.234. Feature contributions show that 1,191 other features collectively decrease the readmission probability by 1%. Notably, a 4-day length of stay significantly reduces the probability by 11%. These contributions combine to yield a predicted probability of 0.006, *f*(*x*) = 0.006.

**Figure A. 2.**
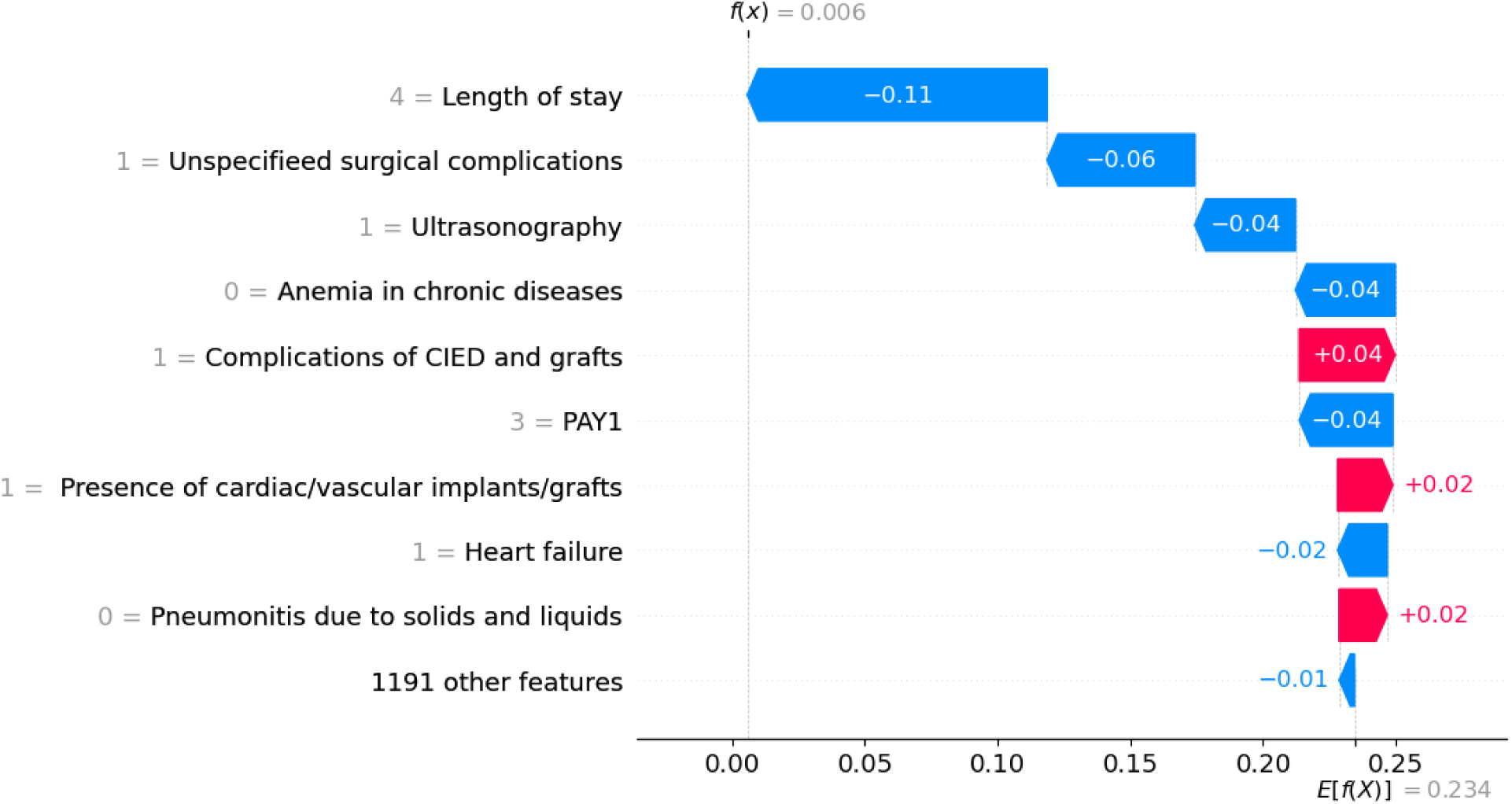
Feature Contributions for Predicting 30-Day Readmission. This figure illustrates the feature contributions for a specific individual who was not readmitted within 30 days. The model predicted a low readmission probability of 0.006, effectively indicating a negligible likelihood of readmission.

## Notes

### Competing Interest Statement

The authors have declared no competing interest.

### Funding Statement

This study did not receive any funding

### Author Declarations

The data were from National readmission database 2019, here is the link to the related website: https://hcup-us.ahrq.gov/db/nation/nrd/Introduction_NRD_2019.jsp#:∼:text=Unweighted%2C%20the%20NRD%20contains%20approximately,national%20readmissions%20for%20all%20payers.

### Summary of Updates

In this version, author affiliations have been updated.

## References

1. Van Diepen S, Katz JN, Albert NM, et al. Contemporary Management of Cardiogenic Shock: A Scientific Statement From the American Heart Association. Circulation. 2017;136(16). doi:10.1161/CIR.0000000000000525

2. Vahdatpour C, Collins D, Goldberg S. Cardiogenic Shock. J Am Heart Assoc. 2019;8(8). doi:10.1161/JAHA.119.011991

3. Samsky MD, Morrow DA, Proudfoot AG, Hochman JS, Thiele H, Rao S V. Cardiogenic Shock After Acute Myocardial Infarction. JAMA. 2021;326(18):1840. doi:10.1001/jama.2021.18323

4. VanDyck TJ, Pinsky MR. Hemodynamic monitoring in cardiogenic shock. Curr Opin Crit Care. 2021;27(4):454–459. doi:10.1097/MCC.0000000000000838

5. Jentzer JC, van Diepen S, Barsness GW, et al. Cardiogenic Shock Classification to Predict Mortality in the Cardiac Intensive Care Unit. J Am Coll Cardiol. 2019;74(17):2117–2128. doi:10.1016/j.jacc.2019.07.077

6. Mahmoud AN, Elgendy IY, Mojadidi MK, et al. Prevalence, Causes, and Predictors of 30-Day Readmissions Following Hospitalization With Acute Myocardial Infarction Complicated By Cardiogenic Shock: Findings From the 2013–2014 National Readmissions Database. J Am Heart Assoc. 2018;7(6). doi:10.1161/JAHA.117.008235

7. Amoateng R, Ahmed I, Lander M. Abstract 14034: 30-day Readmission Rates of Cardiogenic Shock: A Nationwide Analysis of Inpatient Demographics and Clinical Outcomes. Circulation. 2022;146(Suppl_1). doi:10.1161/CIRC.146.SUPPL_1.14034

8. Sud K, Haddadin F, Tsutsui RS, et al. Readmissions in ST-Elevation Myocardial Infarction and Cardiogenic Shock (from Nationwide Readmission Database). Am J Cardiol. 2019;124(12):1841–1850. doi:10.1016/J.AMJCARD.2019.08.048

9. Shah M, Patil S, Patel B, et al. Causes and Predictors of 30-Day Readmission in Patients With Acute Myocardial Infarction and Cardiogenic Shock. Circ Heart Fail. 2018;11(4). doi:10.1161/CIRCHEARTFAILURE.117.004310

10. Ascent of machine learning in medicine. Nat Mater. 2019;18(5):407–407. doi:10.1038/s41563-019-0360-1

11. Rajula HSR, Verlato G, Manchia M, Antonucci N, Fanos V. Comparison of Conventional Statistical Methods with Machine Learning in Medicine: Diagnosis, Drug Development, and Treatment. Medicina (B Aires). 2020;56(9):455. doi:10.3390/medicina56090455

12. Deo RC. Machine Learning in Medicine. Circulation. 2015;132(20):1920–1930. doi:10.1161/CIRCULATIONAHA.115.001593

13. Li S, Hickey GW, Lander MM, Kanwar MK. Artificial Intelligence and Mechanical Circulatory Support. Heart Fail Clin. 2022;18(2):301–309. doi:10.1016/j.hfc.2021.11.005

14. Hu Y, Lui A, Goldstein M, et al. Development and external validation of a dynamic risk score for early prediction of cardiogenic shock in cardiac intensive care units using machine learning. Eur Heart J Acute Cardiovasc Care. Published online March 22, 2024. doi:10.1093/ehjacc/zuae037

15. Zweck E, Kanwar M, Li S, et al. Clinical Course of Patients in Cardiogenic Shock Stratified by Phenotype. JACC Heart Fail. 2023;11(10):1304–1315. doi:10.1016/J.JCHF.2023.05.007

16. Golas SB, Shibahara T, Agboola S, et al. A machine learning model to predict the risk of 30-day readmissions in patients with heart failure: a retrospective analysis of electronic medical records data. BMC Med Inform Decis Mak. 2018;18(1):44. doi:10.1186/s12911-018-0620-z

17. Lin, Tianyang, et al. "A survey of transformers." AI open 3 (2022): 111–132.

18. Gorishniy, Yury, et al. "Revisiting deep learning models for tabular data." Advances in Neural Information Processing Systems 34 (2021): 18932–18943.

19. Lundberg SM, Lee SI. A unified approach to interpreting model predictions. Adv Neural Inf Process Syst. Published online 2017.

